# DNA methylation markers for sensitive detection of circulating tumor DNA in patients with gastroesophageal cancers

**DOI:** 10.1101/2024.03.04.24303699

**Authors:** Nadia Øgaard, Cecilie R. Iden, Sarah Østrup Jensen, Salah Mohammad Mustafa, Emilie Aagaard, Jesper Bertram Bramsen, Lise Barlebo Ahlborn, Jane Preuss Hasselby, Kristoffer Staal Rohrberg, Michael Patrick Achiam, Claus Lindbjerg Andersen, Morten Mau-Sørensen

## Abstract

**Background:** Patients with gastric and gastroesophageal junction adenocarcinomas (G-GEJ AC) face poor outcomes. Thus, sensitive biomarkers for improved clinical management are highly warranted. Detection of circulating tumor DNA (ctDNA) using DNA methylation biomarkers is a highly sensitive approach for cancer detection and management. Here, we explored the potential of a tumor-agnostic test targeting DNA methylation to detect ctDNA in patients with resectable and advanced G-GEJ AC.

**Methods:** Tumor DNA from 29 patients, and plasma cell-free DNA from 17 patients with advanced- and 17 patients with resectable G-GEJ AC, and from 50 healthy controls was analyzed. A tumor-agnostic, digital PCR test – TriMeth - targeting the gastrointestinal cancer-specific methylated genes *C9orf50, KCNQ5*, and *CLIP4*, was performed.

**Results:** TriMeth detected tumor DNA in 29/29 (100%) of the tumor tissue samples. Furthermore, TriMeth detected ctDNA in plasma from 13/17 (76%) of patients with advanced disease, 7/17 (41%) of patients with resectable disease, and in 0/50 (0%) of healthy controls.

**Conclusions:** This study demonstrates that TriMeth may hold potential as a biomarker for identification of ctDNA in patients with G-GEJ AC. The study sets the scene for ongoing larger clinical studies investigating the performance of TriMeth in different clinical settings.

## Introduction

In 2020, gastric and gastroesophageal junction cancers were responsible for more than 1.3 million deaths worldwide, making it the third leading cause of cancer-related death globally [1, 2]. Up to 90% of gastric and gastroesophageal junction (GEJ) cancers are adenocarcinomas. Gastric and GEJ adenocarcinomas (G-GEJ AC) share many of the same clinicopathological traits, with GEJ AC being the most prevalent. Patients with G-GEJ ACs carry a dismal prognosis, and are often diagnosed at an advanced stage [3]. This limits the curative potential of available therapeutic interventions as reflected by the poor 5-year survival rate of 20% across stages [4]. Even patients with resectable disease have a 5-year survival of less than 50% [5]. Hence, there is an urgent need for sensitive biomarkers for surveillance of recurrence and monitoring of treatment benefits in patients with G-GEJ AC.

Circulating tumor DNA (ctDNA) is a surrogate for tumor burden. ctDNA enters the bloodstream when DNA is released from dying tumor cells. ctDNA can be quantified from routinely collected peripheral blood, allowing minimally invasive assessment of ctDNA fragments in the circulation [6, 7]. In recent years, detection of ctDNA through DNA methylation alterations has broken new grounds for cancer diagnostics and surveillance [8, 9, 10]. In contrast to tumor-informed strategies, ctDNA methylation detection has the major advantage of being tumor-agnostic i.e. does not require prior tumor tissue analysis.

We have previously shown that detection of ctDNA using a multiplex digital PCR test targeting the gastrointestinal cancer-specific DNA methylation markers *C9orf50, KCNQ5*, and *CLIP4* (the TriMeth test) is a highly sensitive approach for early detection of colorectal cancer (CRC) and CRC recurrence [9, 11, 12]. If this method can also provide sensitive detection of G-GEJ AC, it could potentially improve the clinical management of G-GEJ AC patients.

Here, we investigated the performance of TriMeth in tissue from patients with resectable G-GEJ AC, and furthermore explored the potential of the TriMeth test to detect ctDNA in plasma from patients with resectable and advanced G-GEJ AC.

## Material and methods

### Study design

This is a retrospective study exploring the performance of the TriMeth test in tumor tissue and plasma from patients with G-GEJ AC. Due to the explorative design, no sample size estimation was performed. A brief study overview is illustrated in Figure 1. A total of 73 study subjects were included; tumor tissue was collected from 29 patients with resectable G-GEJ AC, while plasma was collected form 17 patients with advanced disease, 17 patients with resectable disease, and from 50 healthy individuals (Figure 1). DNA was extracted from all samples and analyzed by the multiplex droplet digital PCR TriMeth test.

**Figure 1.**
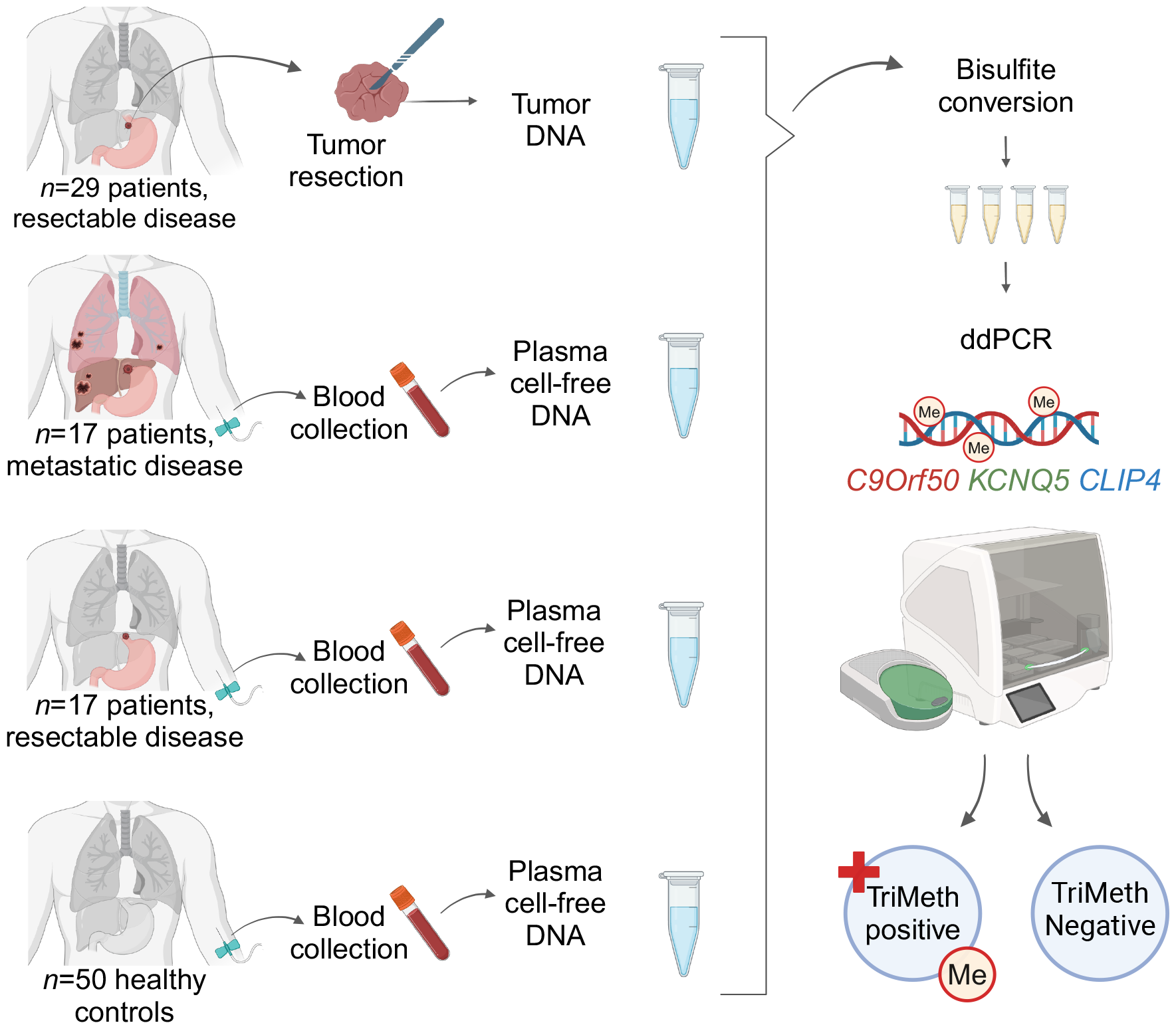
Study design. Tumor DNA from fresh frozen surgical specimens of 29 patients resected for gastric and GEJ adenocarcinomas, and plasma circulating cell-free DNA from 17 patients with metastatic- and 17 patients with resectable gastric and GEJ adenocarcinomas, and from 50 healthy controls were extracted. Tumor DNA and cell-free DNA were treated with Sodium Bisulfite prior to methylation analysis. All samples were analyzed using a DNA methylation-based multiplex digital PCR test termed ‘TriMeth’, which targets the promoter regions of the gastrointestinal cancer-specific methylated genes *C9orf50, KCNQ5*, and *CLIP4*. Samples were classified as ‘TriMeth positive’ if at least 2/3 markers were positive with >1 droplet. ddPCR: droplet digital PCR. Illustration created using BioRender

**Figure 2.**
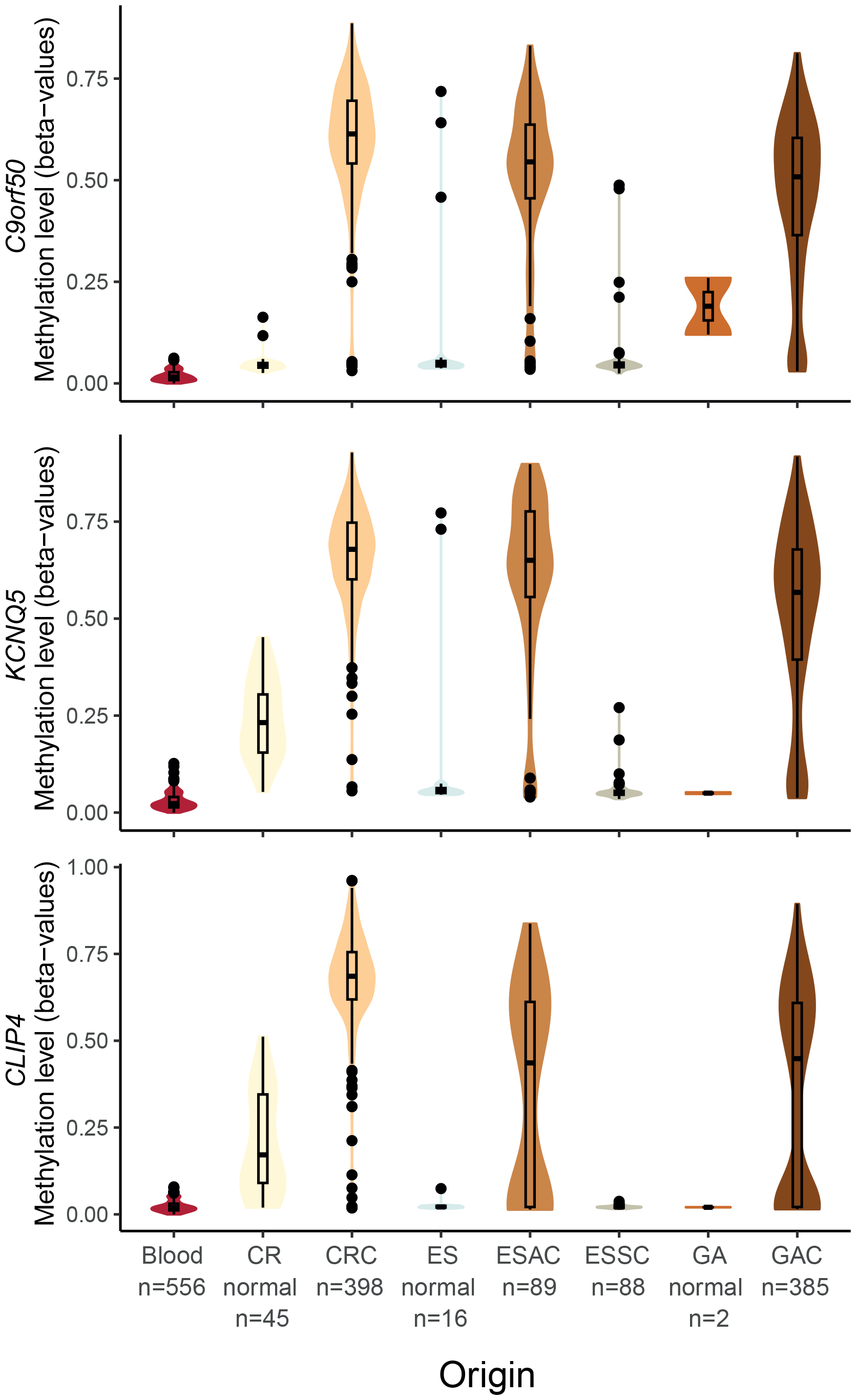
Methylation levels of TriMeth markers across gastrointestinal cancers. Methylation levels of C9orf50, KCNQ5, and CLIP4 in tissue samples from: 45 normal colorectal mucosa (CR normal), 398 colorectal cancers (CRC), 16 normal esophagus (ES normal), 89 esophageal adenocarcinomas (EAC), 88 esophageal squamous cell carcinomas (ESCC), 2 normal gastric tissue (GA normal), 385 gastric adenocarcinomas (GAC), and 556 blood cell samples. Beta-values (the ratio of the methylated probe intensity and the overall probe intensity of probes covering the candidate CpG-sites) were acquired from Infinium HumanMethylation450K BeadChip array data generated by the Cancer Genome Atlas (TCGA) project [samples barcodes can be found in Supplemental Table S1]. Solid horizontal bar (median); open vertical box (interquartile range); solid vertical line (1.5 x interquartile range); colored area (distribution of data points); solid dots (outliers). Illustration created using Adobe

### Patients and samples

Fresh frozen surgical tumor specimens were collected from 29 patients undergoing surgery for G-GEJ AC cancer between 2014 and 2016 at the Department of Surgery and Transplantation at Copenhagen University Hospital, Rigshospitalet, Denmark. All 29 patients received curatively intended, standard-of-care treatment i.e., perioperative chemotherapy and surgical resection. Plasma samples were collected from a total of 84 individuals, including 17 patients with advanced G-GEJ AC, 17 patients with resectable G-GEJ AC, and 50 healthy individuals.

The 17 patients with advanced G-GEJ AC were referred to The Phase 1 Unit at The Department of Oncology at Copenhagen University Hospital, Rigshospitalet, Denmark. These patients were included in the Copenhagen Prospective Personalized Oncology (CoPPO) study between 2018 and 2019, with the aim to allocate patients with exhausted treatment options to early clinical trials of targeted therapies based on genomic profiling [13, 14]. Plasma was collected at the time of referral to The Phase 1 Unit.

The 17 patients with resectable G-GEJ AC, were included in the prospective study Clinical Utility of Circulating tumor DNA (CURE) [15] at The Department of Oncology at Copenhagen University Hospital, Rigshospitalet, Denmark. These patients received curatively intended treatment with perioperative chemotherapy with eight cycles of docetaxel, oxaliplatin, and fluorouracil/leucovorin (FLOT) and surgical resection according to national guidelines [16, 17]. Plasma was collected prior to oncological treatment.

As negative controls, 50 non-cancer individuals from the Danish national CRC screening program were included [18]. These all had a plasma sample collected in relation to a positive fecal immunochemical test during the period of 2014-2016. All individuals showed a clean colon on the subsequent colonoscopy, and none of them had any comorbidities or previous diagnoses of cancer.

The Ethics Committee of the Capital Region Denmark approved the collection and use of biological samples [j. nos. H-16046103; H-4-2013-050; H-2-2013-101; H-19076846; H-20033168] and the capture of clinical data from patient journals [j. nos. H-21052982; H-1300530; H-16046103]. Informed consent was obtained from all participating patients. The Danish Data Protection Agency [j. nos. HVH-2013-022/2007-58-0015; P-2019-701] approved the study.

### Methylation array data

DNA methylation data generated by The Cancer Genome Atlas (TCGA) project including esophagus cancer (“ESCA”), stomach adenocarcinoma (“STAD”), and colorectal cancer (“COAD+READ”) samples were acquired from the GDC data portal (v1.30.0; Data Release 34.0, March 20th 2023) using the following filters: Data Type=“Methylation Beta Value”, Platform= “illumina human methylation 450”, Workflow Type= “SeSAMe Methylation Beta Estimation”, “Data Format”, Samples Is FFPE=“FALSE” [19]. Only primary tumor and normal samples were included. TCGA File IDs are available in Supplemental Table S1. The histological type of the tumors was previously defined by Liu et al. [20]. Methylation array data from 556 blood cell samples was downloaded and processed as previously published by Jensen et al. [11]. Beta-values (the ratio of the methylated probe intensity and the overall intensity) for each sample were evaluated for each of the TriMeth marker candidate CpG sites (*C9orf50* probe: cg14015706, *CLIP4* probe: cg08808128, *KCNQ5* probe: cg24687051), which were identified by Jensen et al. [11].

### DNA methylation analysis (TriMeth)

The three TriMeth markers were identified and selected in a previous study employing DNA methylation array data from >5000 samples for bioinformatics marker discovery, described in detail by Jensen et al [11]. Here, the TriMeth marker assays were developed and tested and validated in tissue and plasma from colorectal cancer patients. In the present study, TriMeth analysis was performed according to previous procedures [11, 12] using methylation-specific droplet digital PCR assays (ddPCR) targeting the promoter regions of *C9orf50, CLIP4*, and *KCNQ5*, and a quantification assay (CF assay). All TriMeth assays are listed in Supplemental Table S2. Details on sodium bisulfite conversion, and droplet digital PCR can be found in the Supplementary Materials and Methods. Each analysis included a positive control (5 ng of fully methylated DNA, Nordic Biosite), a negative control (66 ng of fully unmethylated DNA, Nordic Biosite), and a non-template control. For each plate, a threshold was determined by evaluation of the positive and negative controls as previously described [11, 12].

### Data analysis

A TriMeth score was assigned to each sample using a pre-defined cut-off classifying a sample as ‘positive’ if >1 positive droplet was recorded for 2 out of 3 TriMeth markers [11, 12].

For the tissue samples [Figure 3]: the input DNA (in Genome equivalents (GE)) for the TriMeth analysis was assessed for each sample by a cytosine free DNA quantification assay (CF) which provides a measure of the total number of DNA copies in the analysis quantification digital PCR assay [Supplementary Material and Methods]. For each marker we calculated the methylated allele frequency (AF), as ‘the number of methylated DNA copies / the input DNA’. The mean methylation AF (mean meth AF) was calculated as the mean of the three marker’s methylated AFs. This measure illustrates how much of the analyzed DNA on average was methylated in the given sample [Supplemental Table S3]. The relative methylation levels (calculated as the methylation signal for an individual marker / sum of methylation signal for all three markers) [Supplemental Table S3] provides a measure of the relative abundance of methylation at the three markers. Spearman analysis was performed to estimate the correlation between histologically evaluated tumor cell density and methylated AF assessed by TriMeth.

**Figure 3.**
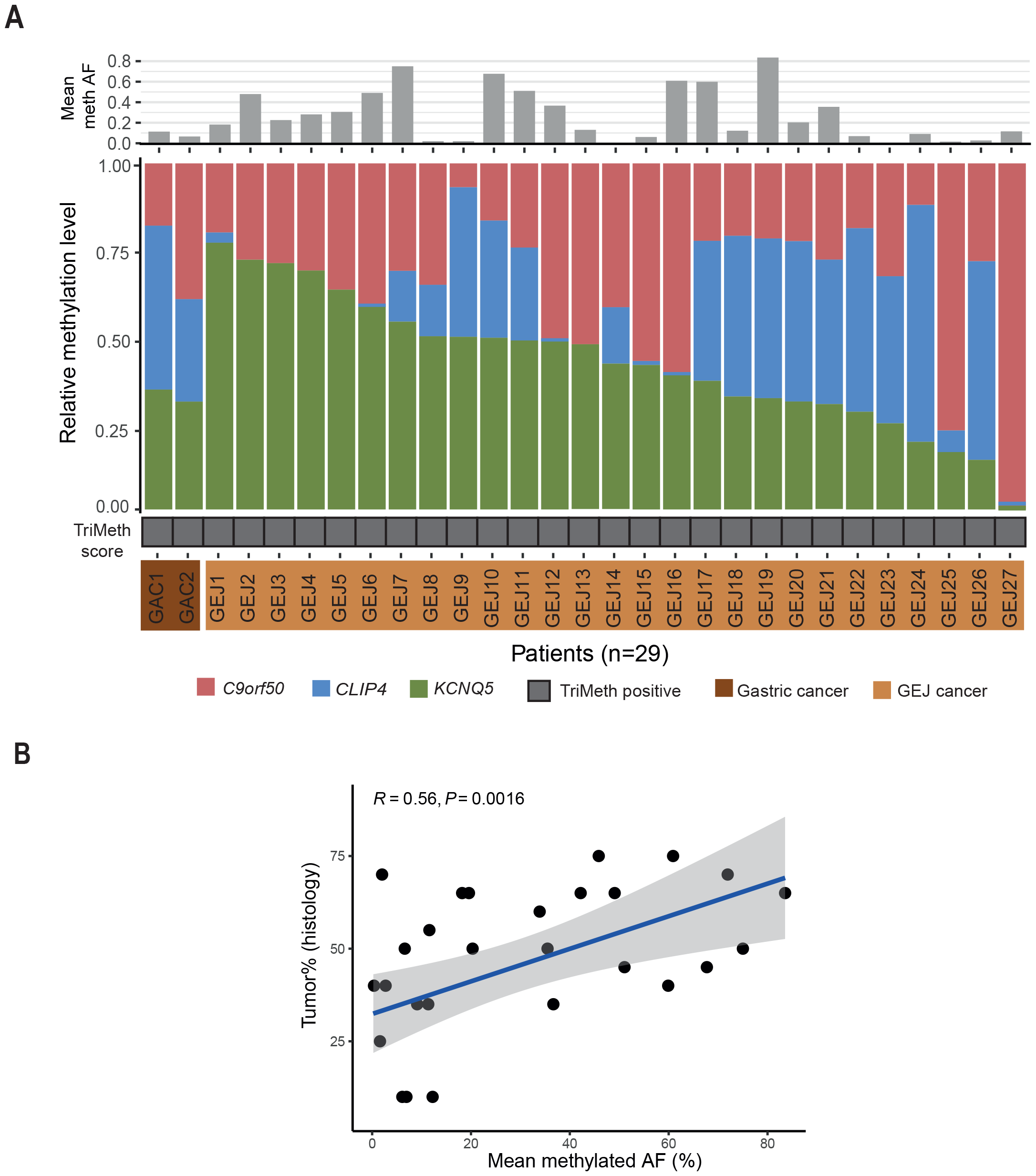
TriMeth methylation in gastric and GEJ tumor tissue. **a**) For each tumor sample (n=29) analyzed by TriMeth the following are shown (from top): mean methylated allele frequency (AF) (calculated as the mean AF of the three markers); relative methylation level (calculated as the methylation signal for an individual marker / sum of methylation signal for all three markers); marker status (positive or negative/not detected). **b**) Correlation of mean methylated AF by TriMeth and the histologically estimated tumor cell density (tumor%) in the sample, assessed by Spearman. Illustration created using Adobe

For the plasma samples [Figure 4]: the methylation signal for each marker was calculated as the methylated copies detected by the marker divided by the analyzed plasma volume, and log-transformed for improved visualization. The non-logged (the absolute) methylated ctDNA copies detected by each marker and all calculations for the plasma samples are presented in Supplemental Table S4.

**Figure 4.**
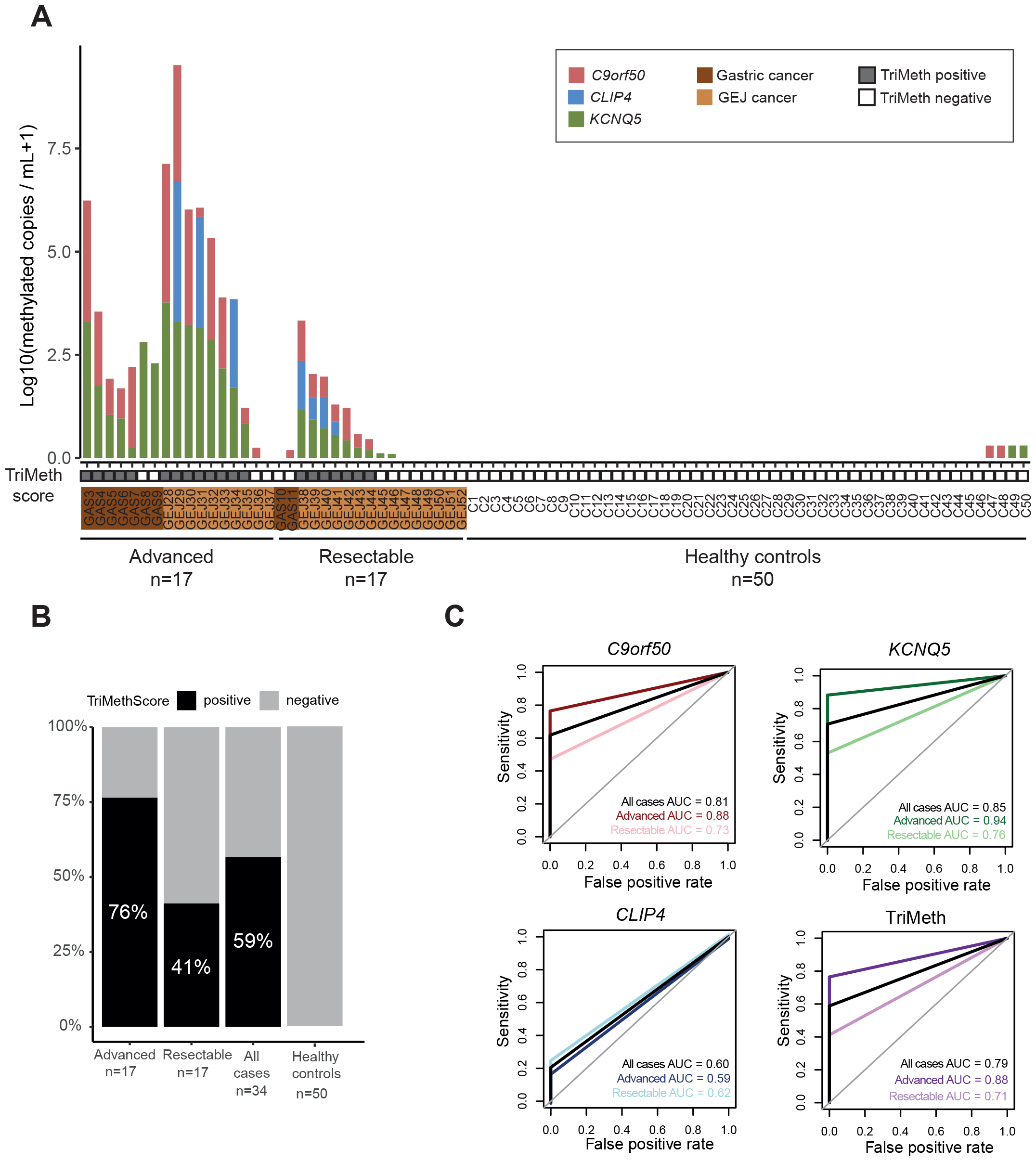
TriMeth for ctDNA detection in plasma from patients with gastric and GEJ cancer. **a**) Methylation levels of *C9orf50, CLIP4*, and *KCNQ5* and TriMeth score in plasma from patients with advanced or resectable gastric and GEJ cancer, and healthy individuals. **b**) Fraction of individuals with TriMeth positive or negative score (TriMeth sensitivity). c) ROC curves and AUCs based on ctDNA detection. ROC: receiver operating characteristic, AUC: area under the curve. Illustration created using Adobe

The predictive accuracy of the individual markers and of the TriMeth test was estimated by receiver operating characteristic (ROC) analysis using the R package ROCR. All analyses were performed using R software version 4.1.1 and 4.2.2.

Additional descriptions of the applied methods can be found in Supplementary Material and Methods.

## Results

In this study, we investigated the performance of three DNA methylation markers, TriMeth, in tissue and plasma from patients with G-GEJ AC. An overview of the study workflow is presented in Figure 1. We included 29 patients with resectable G-GEJ AC for tumor tissue analysis. All 29 patients were treated with standard-of-care perioperative chemotherapy and resection. Their median age was 71 years, 82.8% were male, and for 93.1% of the patients, the tumor was localized in the lower esophagus or GEJ. For plasma analysis, we included 17 patients with resectable G-GEJ AC. All 17 patients received curatively intended perioperative chemotherapy and resection. Their median age was 68 years, 70.6% were male, and for 88.2% of the patients the tumor was localized in the lower esophagus or GEJ. For plasma analysis, 17 patients with advanced G-GEJ AC were also included. Their median age was 60 years, 82.4% were male, and 58.8% of their tumors were located in the lower esophagus or the GEJ. Finally, we included plasma from 50 non-cancer control individuals. The median age of the healthy controls was 62 years, and 76.0% were male. Patient characteristics are summarized in Table 1.

**Table 1.**
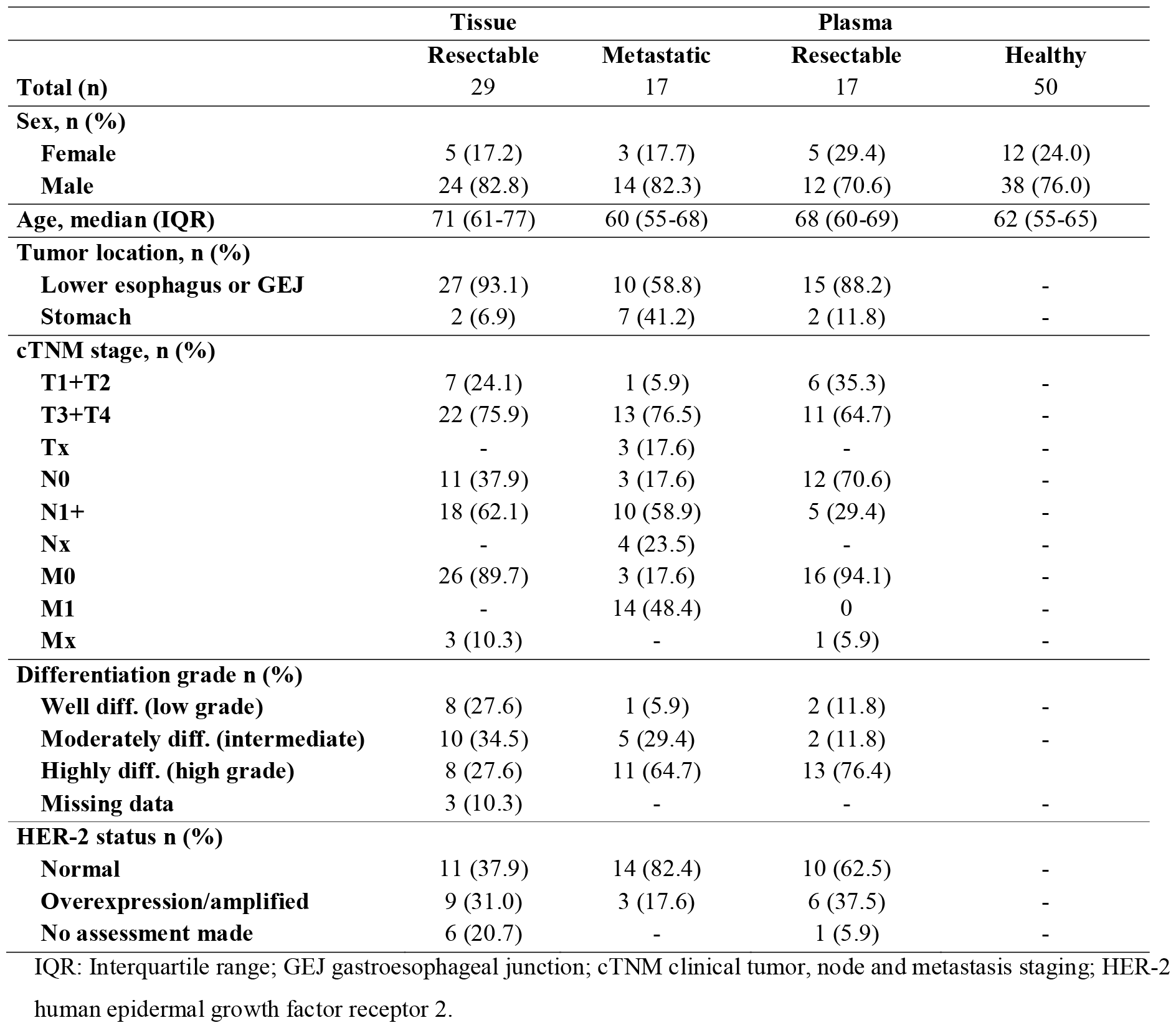
Patient characteristics and demographics.

### Methylation levels of TriMeth markers across gastrointestinal cancers

To gain insight into the overall methylation levels of the TriMeth markers across gastrointestinal cancers, we performed an in-silico assessment of the marker methylation levels using publicly available DNA methylation array data [Figure 2[. All three markers were negligibly methylated in peripheral blood mononuclear cells, and were minimally methylated in most normal epithelial samples from esophageal, gastric, and colorectal tissues [Figure 2]. All markers were highly methylated in CRC samples (medians of 0.61 for *C9orf50*, 0.68 for *KCNQ5*, and 0.69 for *CLIP4*). The esophageal cancer samples were divided into the histological subtypes adenocarcinoma (EAC) and squamous cell carcinoma (ESCC). Notably, the methylation levels of the TriMeth markers varied greatly between the two subtypes. In ESCC samples, the markers showed no or minimal methylation, comparable to the methylation of the blood cell samples, whereas all three markers were highly methylated in the majority of EAC samples. In gastric adenocarcinoma (GAC) samples, all three markers were moderately methylated (medians of 0.51 for *C9orf50*, 0.57 for *KCNQ5*, and 0.45 for *CLIP4*). For the *CLIP4* marker, around 25% of the EAC and GAC samples showed no methylation [Figure 2]. Since the TriMeth marker regions were unmethylated in ESCC samples, only gastric and esophageal cancer patients with an AC subtype were included in TriMeth analyses of tissue and plasma.

### TriMeth methylation in gastric and GEJ tumor tissue

We analyzed the performance of the TriMeth markers in tumor DNA from 29 patients with resectable G-GEJ AC. Detailed TriMeth data of the 29 tumor samples is presented in Online Resource 2, Table S3. A median of 6,050 GE (interquartile range (IQR): 5,675 - 6,424) of bisulfite converted tumor DNA was used as input for TriMeth analysis [Supplemental Table S3 and Figure 3]. Methylated DNA was detected by TriMeth in all 29/29 (100%) of the tumor samples (Figure 3a). The relative methylation level of the individual TriMeth markers was plotted to investigate the relative abundance of methylation of the three markers in the tissues. The *KCNQ5* and *C9orf50* markers showed the most persistent contribution to the TriMeth methylation signal (*KCNQ5* median: 0.44, IQR: 0.33-0.52, and *C9orf50* median: 0.28, IQR: 0.21-0.40), whereas the *CLIP4* relative methylation level varied greatly between the samples [Figure 3a]. In 10/29 samples, *CLIP4* showed the highest methylation level of the three markers, and in 5/29 samples no *CLIP4* methylation was detected. The mean methylated AF assessed by TriMeth ranged 0.4% to 83.6% [Figure 3a]. The mean methylated AF was moderately correlated to the histologically estimated tumor cell density (tumor%) (R = 0.56, p= 0.0016, Figure 3b). In summary, all investigated tumor specimens were TriMeth positive.

### TriMeth ctDNA detection in plasma from patients with G-GEJ AC

Next, we aimed to assess the performance of TriMeth for ctDNA detection in plasma from G-GEJ AC patients. For this analysis we included 17 patients with advanced G-GEJ AC, 17 patients with resectable G-GEJ AC, and 50 healthy controls. A plasma volume of 4 mL was used as input for the TriMeth analysis with a median input of 1,884 GE (IQR: 1,621-2.293) of bisulfite converted cfDNA for the controls and a median input of 4,158 GE (IQR: 2,136-5,862) of bisulfite converted cfDNA for the cases.

The mean methylated ctDNA copies detected by the TriMeth markers ranged from 0.5 GE/mL to 2,713 GE/mL [Supplemental Table S4]. In total, 13 out of 17 (76.5%) plasma samples from the patients with advanced G-GEJ AC were scored as TriMeth positive [Figures 4a-b and Supplemental Figure S1]. In patients with resectable G-GEJ AC, 7 out of 17 (41.2 %) samples were TriMeth positive [Figures 4a-b and Supplemental Figure S1]. Six TriMeth-negative cases were positive for one of the three markers. For four of these, the methylation levels were low, but for two samples (GAC8 and GAC9) > 500 methylated ctDNA copies were detected by *KCNQ5* [Figure 4a and Supplemental Table S3]. The *CLIP4* marker was blank in all plasma samples from patients with gastric cancer. Four out of fifty plasma samples from the healthy individuals were positive for 1/3 TriMeth markers, accordingly, all healthy controls were scored as TriMeth negative [Figures 4a and b].

Compared to *CLIP4*, the performance of *C9orf50* and *KCNQ5* was higher across all patients regardless of disease type and stage with area under the curve (AUC)s of 0.73 - 0.88 and 0.76 - 0.94 [Figure 4c]. The AUC of TriMeth for all cases was 0.79. Detailed TriMeth data of plasma samples is presented in Supplemental Table S4.

In summary, the TriMeth markers detected ctDNA in plasma from patients with G-GEJ AC with high sensitivity, whereas all plasma samples from the healthy controls were negative.

## Discussion

In the last decades, multiple tissue-based biomarkers enabling personalized management of solid cancers have been developed. Recently, blood-based biomarkers, such as ctDNA, have shown promising potential to take personalized cancer management to the next level. ctDNA-based biomarkers have particularly promising potential in clinical settings such as detection of recurrence or minimal residual disease and selection and monitoring of targeted therapy [21]. So far, no ctDNA-based biomarkers have been identified and validated for routine clinical use in patients with G-GEJ AC [22]. Here, we investigated the performance of previously developed tumor-agnostic test, TriMeth, targeting gastrointestinal DNA hypermethylation in tissue and plasma from G-GEJ AC patients and demonstrate that TriMeth may hold potential for robust identification of ctDNA in patients with G-GEJ AC.

To investigate the TriMeth marker methylation levels across gastrointestinal cancers, we analyzed 450K methylation array data, and showed that the TriMeth markers were highly methylated in CRC, EAC, and GAC, whereas the ESCC samples were unmethylated. Therefore, only patients with G-GEJ AC were included in TriMeth analyses of tissue and plasma, and it should be noted that TriMeth may not be suitable for ctDNA detection in patients with ESCC.

When we investigated TriMeth in G-GEJ AC tumor tissue, all samples (29/29, 100%) were classified as TriMeth-positive. While *C9orf50* and *KCNQ5* were positive in all 29/29 samples and these markers had the strongest contribution to the overall TriMeth methylation signal, the *CLIP4* marker was more infrequently methylated [Figure 3]. This is in correlation with the 450K array data showing that ∼25% of the GAC samples had no *CLIP4* methylation [Figure 2]. The mean methylated AFs measured by TriMeth in the tissue samples varied greatly between the patients, from 0.4% to 83%, and were generally lower compared to the levels previously reported in CRC tissue [11]. However, this observation is coherent with the methylation levels from the array data [Figure 2], indicating a slightly lower methylation of all three markers in G-GEJ AC samples compared to the CRC samples.

In plasma, TriMeth detected ctDNA in 13/17 (76%) patients with advanced G-GEJ AC. Interestingly, two of the TriMeth-negative patients (GAC8 and GAC9) were positive with more than 200 copies for *KCNQ5*. In previous published and unpublished work, a false positive *KCNQ5* signal >5 copies have never been observed (in >1,000 analyzed plasma samples from healthy controls) [11, 12]. Hence, considering these two samples as positive, the sensitivity of TriMeth in patients with advanced G-GEJ AC would be 88% (15/17).

*KCNQ5* and *C9orf50* contributed consistently to the TriMeth positivity in plasma (AUCs of 0.85 and 0.81 for all cases), whereas the combined AUC of the TriMeth test was 0.79. These results confirmed the findings of a recent study by Li et al. [23] who tested the *CLIP4, C9orf50*, and *KCNQ5* markers in gastric cancer plasma cohorts by qPCR and reported AUCs of 0.78-0.85.

This research sets the scene for studies unravelling the clinical utility of this test within different clinical settings of gastric- and gastroesophageal junction cancer management, such as detection of minimal residual disease detection after surgery, recurrence surveillance, guidance in treatment selection, and monitoring of treatment effects, which may all contribute to improved patient outcomes.

Although having intriguing potential, a few limitations the study should be acknowledged. Firstly, the study had a small sample size, and validation of TriMeth in larger cohorts are needed to confirm the potential of TriMeth in G-GEJ AC. Secondly, tissue and plasma were collected form two independent cohorts, and therefore no matched samples were available, limiting the possibility to investigate concurrent methylation of matched tissue and plasma. Thirdly, the healthy controls had endoscopic examination of the colon and rectum and not of the upper gastrointestinal tract. Finally, it should be noted that the TriMeth markers and the TriMeth 2/3 scoring algorithm were developed to be sensitive towards early detection of CRC. Therefore, it is likely that the sensitivity of the TriMeth test towards G-GEJ AC could be improved by adjusting the TriMeth scoring algorithm to be less conservative or by including methylation markers more sensitive towards G-GEJ AC.

In conclusion, the TriMeth assay detected ctDNA in patients with G-GEJ AC with high sensitivity, elucidating a potential of TriMeth for early detection and detection of recurrence in the clinical management of G-GEJ AC patients. Larger clinical studies are needed to demonstrate the utility of TriMeth in different clinical settings during the disease course of G-GEJ AC patients.

## Data Availability

The original data presented in the study are included in the Supplemental Tables S3-S4. Further inquiries can be directed to the corresponding author.

## Acknowledgements

We would like to thank all patients as well as participants in the Danish national CRC screening program, for contribution of clinical material. We also thank The Departments of Oncology, Clinical Biochemistry, Pathology Surgery and Transplantation and The Phase 1 Unit at Copenhagen University Hospital, Denmark for patient inclusion and management, and The Department of Molecular medicine at Aarhus University Hospital for sample processing and analysis.

The results published here are partly based upon data generated by the TCGA Research Network: https://www.cancer.gov/tcga.

