## Supplementary material for "DNA methylation markers for sensitive detection of circulating tumor DNA in patients with gastroesophageal cancers"

**Tissue and plasma processing and DNA isolation**

Using a cryo-microtome, 27 tissue sections were cut from fresh frozen tissue specimens (5 µm thickness). The first and the last slide were stained with hematoxylin and eosin, and the tumor cell density was histologically estimated in these two slides. The mean tumor cell density of the two slides were registered, as the tumor% of the tumor biopsy. From the remaining 25 tissue sections DNA was extracted using the Gentra Puregene Tissue Kit (Qiagen), following manufacturer’s instructions. The purified DNA was subsequently quantified by Qubit (Termo Fisher Scientific).

**Droplet digital PCR (ddPCR)**

All ddPCR experiments were conducted according to the Minimum Information for Publication of Quantitative Digital PCR Experiments (dMIQE) guidelines 2020 (Suppl. Table S5) (1). All samples were analyzed on a QX200 Droplet Digital PCR system (Bio-Rad) in compliance with the manufacturer’s specifications. Each analysis included positive, negative, and no-template controls. The sample reaction mix included 2-8 µL template DNA, 18 pmol forward primers, 18 pmol reverse primers, 5 pmol probes, 11 µL 2Xsupermix for Probes (Bio-Rad), and nuclease-free water to a final reaction volume of 22 µL. On average 17,786 droplets (interquartile range (IQR): 16,934-18,270) were generated for each sample on the QX200 AutoDG Droplet Generator (Bio-Rad). After droplet generation, samples were amplified by PCR on a S1000 Thermal cycler (Bio-Rad) with the program: 95 °C for 10 minutes, 45 cycles of 95 °C for 30 seconds and 56 °C for one minute, and one final cycle of 98 °C for 10 minutes. PCR products were stored at 4°C for up to 12 hours before they were analyzed on a QX200 reader (Bio-Rad). The Quantasoft v1.7 software (Bio-Rad) was used for analysis of ddPCR data.

**DNA quantification and quality control**

cfDNA purification efficiency and lymphocyte DNA contamination was assessed prior to bisulfite conversion using ddPCR as previously described (2). Primer and probe sequences are listed in Suppl. Table S2. In brief, a fixed amount of soybean CPP1 DNA fragments were added to each plasma sample before cfDNA extraction. Purification efficiency was calculated as the percent recovery of CPP1 fragments after cfDNA extraction (assed by the CPP1 assay). Lymphocyte DNA contamination
