## Supplemental Figure for "DNA methylation markers for sensitive detection of circulating tumor DNA in patients with gastroesophageal cancers"

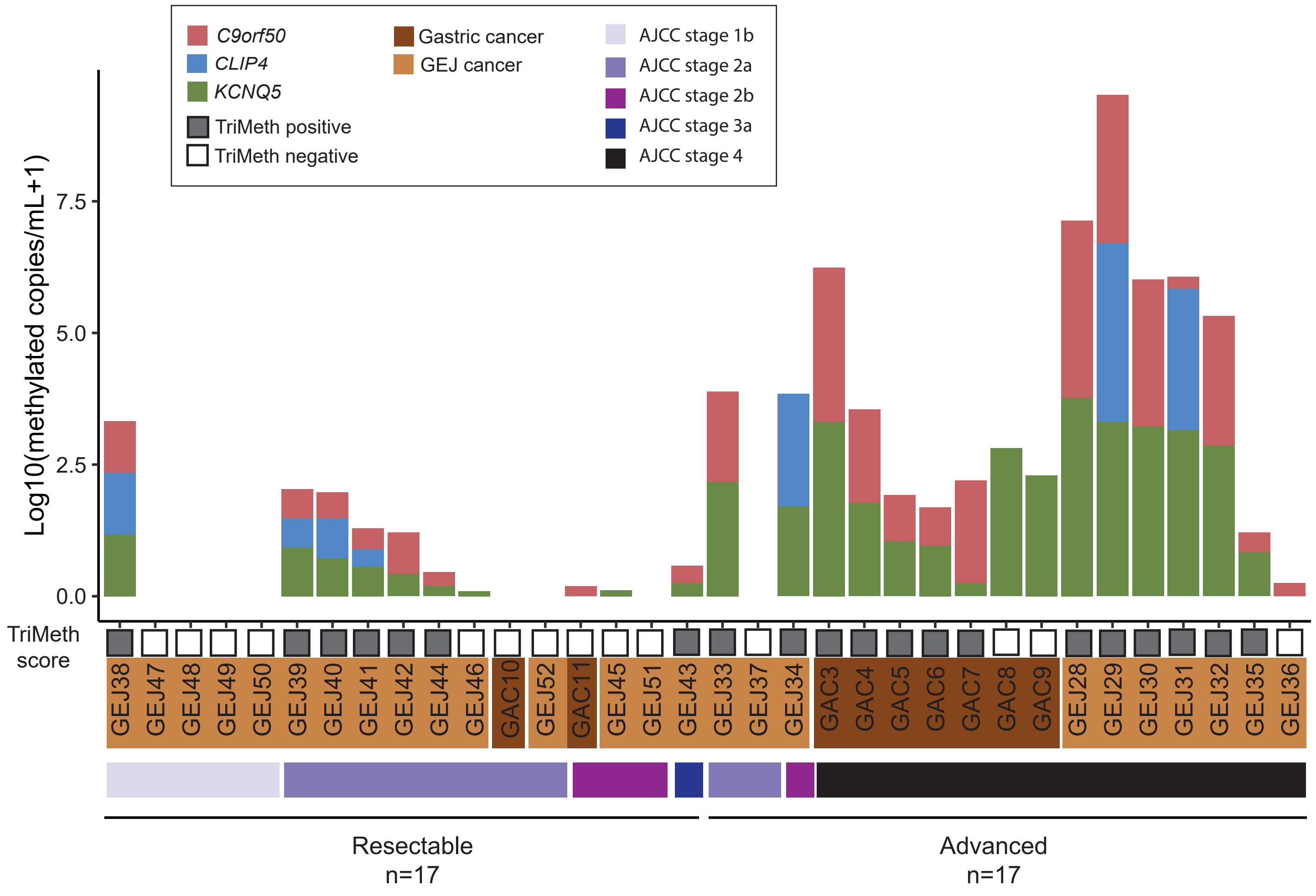


**Supplemental Figure S1**: TriMeth for ctDNA detection in plasma from patients with gastric and GEJ cancer. Methylation levels of *C9orf50*, *CLIP4*, and *KCNQ5* and TriMeth score in plasma from patients with advanced or resectable gastric and GEJ cancer stratified for AJCC stage. AJCC: American joint comitee on cancer.
